# Upper-room ultraviolet air disinfection might help to reduce COVID-19 transmission in buildings

**DOI:** 10.1101/2020.06.12.20129254

**Authors:** Clive B. Beggs, Eldad J. Avital

**Affiliations:** Carnegie School of Sport, Leeds Beckett University, Leeds, UK; School of Engineering and Materials Science, Queen Mary University of London, London, UK

**Keywords:** COVID-19, SARS-CoV-2, ultraviolet, air transmission mitigation, upper room UV-C, presence of humans

## Abstract

As the world economies get out of the lockdown imposed by the COVID-19 pandemic, there is an urgent need to assess the suitability of known technologies to mitigate COVID-19 transmission in confined spaces such as buildings. This feasibility study looks at the method of upper-room ultraviolet (UV) air disinfection that has already proven its efficacy in preventing the transmission of airborne diseases such as measles and tuberculosis.

Using published data from various sources it is shown that the SARS-CoV-2 virus, which causes COVID-19, is highly likely to be susceptible to UV damage while suspended in air irradiated by UV-C at levels that are acceptable and safe for upper-room applications. This is while humans are present in the room. Both the expected and worst-case scenarios are investigated to show the efficacy of the upper-room UV-C approach to reduce COVID-19 air transmission in a confined space with moderate but sufficient height. Discussion is given on the methods of analysis and the differences between virus susceptibility to UV-C when aerosolised or in liquid or on a surface.

## 1. Introduction

Since the emergence of COVID-19 in January 2020 there has been considerable interest in the use of ultraviolet (UV) light to disinfect blood plasma [1-3], equipment [4-7] and air [8], in the hope that this might reduce transmission of the disease. In particular, upper-room ultraviolet germicidal irradiation (UVGI), a technology that disinfects room air, has been muted as a potential intervention that might prove effective against COVID-19 [8-10]. Upper-room UVGI utilizes UV-C light at wavelengths close to 254 nm to create an irradiation field above the heads of room occupants (Figure 1) that disinfects aerosolised bacteria and viruses suspended in the air [11-13]. Because UV-C light is harmful to humans, such systems utilize baffles that obscure the UV lamps from eyesight so that room occupants are safe. As such, upper-room UVGI is a well-established technology [14, 15] that has proven effective as a public health intervention to prevent the spread of airborne diseases such as measles [16] and tuberculosis (TB) [17, 18] in buildings.

**Figure 1.**
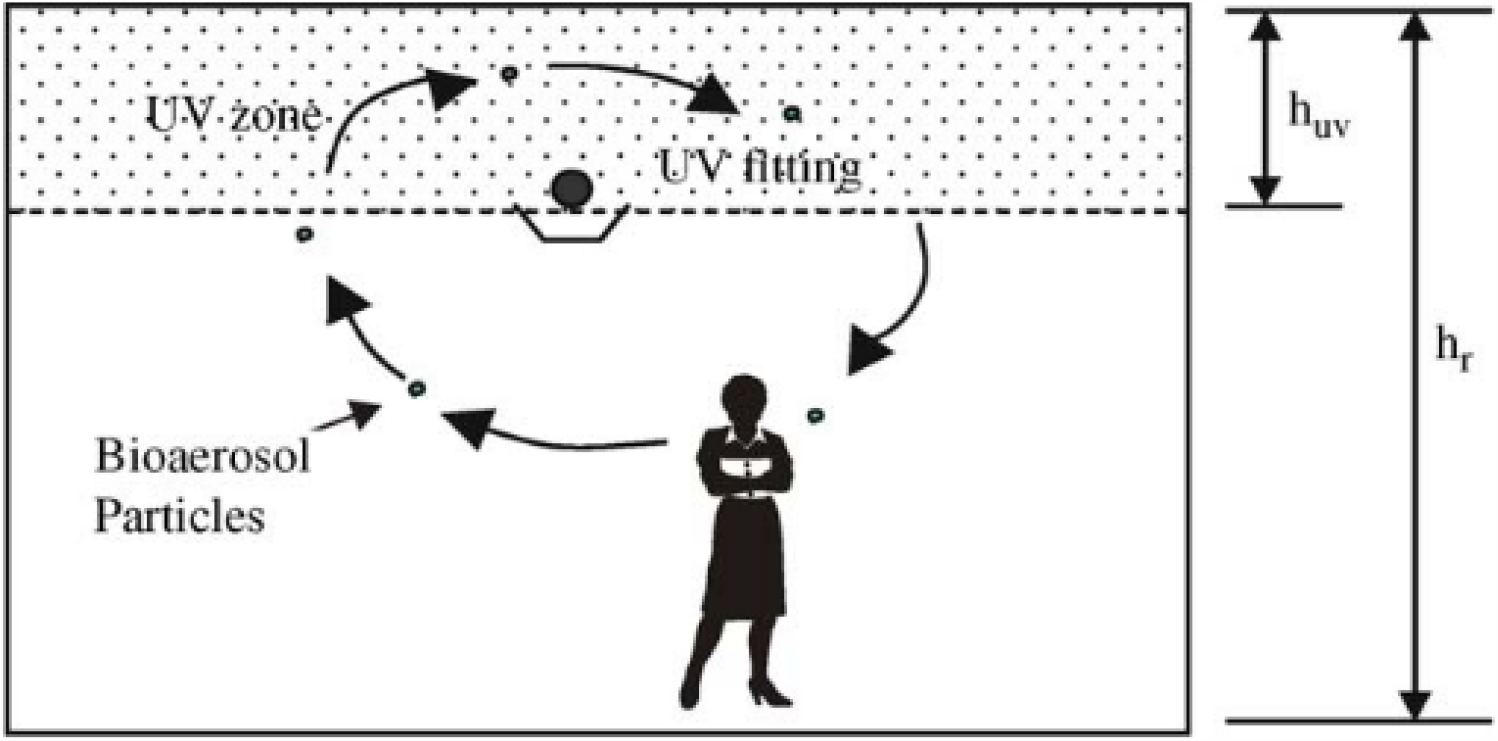
An upper-room UVGI installation.

Given that COVID-19 can be transmitted by the inhalation of aerosolised respiratory droplets containing the SARS-CoV-2 virus [8, 19, 20], and that several studies have recovered viral RNA from hospital air samples [21-24], there is reason to believe that upper-room UVGI might be effective at ‘killing’ (inactivating) SARS-CoV-2 virions in the air, thus reducing the transmission of COVID-19 in buildings and other enclosed spaces. However, this presupposes that the technology is capable of delivering irradiation doses high enough to inactivate SARS-CoV-2 virions in respiratory droplets suspended in the air, something that has not yet been proven. Given this and the urgent need to develop interventions to break the chain of infection associated with COVID-19, we designed the short feasibility study reported here with the aim of evaluating whether or not upper-room UVGI might be an effective intervention against COVID-19.

## 2. Methods

### 2.1 Theory

At any point in time the amount of viral inactivation (disinfection) achieved for a given UV radiant flux (irradiance) can be described using the following first order decay equation [25]: 

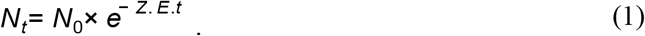

Where: *N*_*0*_ and *N*_*t*_ are the number of viable viral particles (virions) at time zero and *t* seconds respectively; *Z* is the UV susceptibility constant for the virus (m^2^/J); *E* is the radiant (irradiation) flux (W/m^2^); and *t* is time in seconds.

The UV irradiation dose received by the virus is simply: 

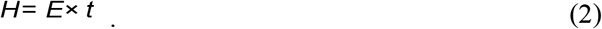

Where: *H* is the observed UV irradiation dose (J/m^2^).

By combining equations 1 and 2, and rearranging we can obtain a value for *Z:* 

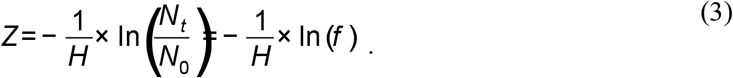

Where: *f* is the survival fraction.

Because the relationship between the UV dose and the natural logarithm of the survival fraction is broadly linear for most viral species, it means that the behaviour of any given virus exposed to UV-C light can be succinctly described by the value of *Z*, irrespective of the actual UV dose applied. As such, for any given viral species, if the value of *Z* is known, then it should be possible to predict with reasonable accuracy how the virus will behave when exposed to a given UV-C dose in any context. Microbes that exhibit larger *Z* values are more susceptible to UV damage, whereas those with small *Z* values are more difficult to inactivate.

UV inactivation plots for most viral species tend to be straight lines, although some might exhibit a curve [26]. Notwithstanding this, the model described in equation 1 is still a good approximation for most viral species [25] up until the point where the ‘target’ becomes saturated with UV photons. At this point, because all the virions have already been inactivated, increasing the UV dose further has no effect and so the linear relationship between UV dose and the log reduction become decoupled, with the result that the *Z* value no long applies.

Instead of quantifying UV inactivation in terms of survival fraction, many researchers, particularly those working in biology, describe the reduction in the microbial count in terms of log reduction, which can be converted to survival fraction as follows: 

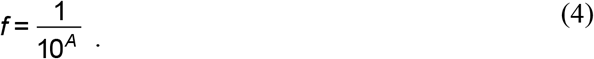

Where: *A* is the log_10_ reduction in the number of viable virions.

Specifically, with regard to upper-room UVGI, once the *Z* value has been obtained for the target microbe, it is then possible to determine the irradiation flux required to disinfect it, using the methodology described in Beggs and Sleigh [11]. This method makes the assumption that the room air is well mixed, which is a reasonable approximation for most applications [11]. If this is the case, then the average particle residence time, *t*_*res*_, (in seconds) in the room space will be: 

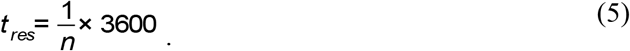

Where: *n* is the room ventilation rate in air changes per hour (AC/h).

From equation 5 it can be approximated that the average particle residence time in the upper-room UV field, *t*_*uv*_, (in seconds) will be: 

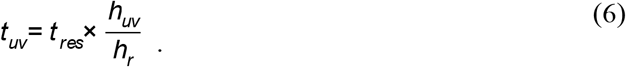

Where: *h*_*r*_ is the floor-to-ceiling height (m), and *h*_*uv*_ is the depth of the upper-room UV zone (m).

Because *Z* values are often determined experimentally using microbes suspended in liquids or on surfaces, it may be necessary to adjust the *Z* value for use with upper-room UVGI systems [12, 27], as follows: 

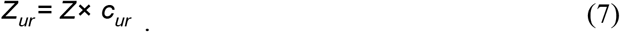

Where: *Z*_*ur*_ is the effective upper-room *Z* value (m^2^/J), and *c*_*ur*_ is a correction coefficient.

So if we assume that the air in a room is well mixed, by combining equations 2, 3 and 6 it is possible to compute the average irradiation flux, *E*_*r*_, that is required to achieve a desired survival fraction, *f*_r_: 

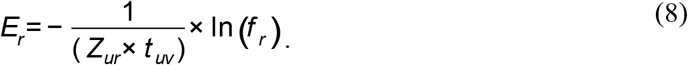

Alternatively, the disinfection achieved by an upper-room UVGI system can be thought of as being equivalent to additional air changes in the room space. In this scenario, the UV rate constant, *k*_*uv*_, which can be thought of as the equivalent air change rate per second, can be determined using [12]: 

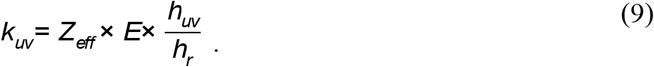

So in a ventilated room in which contamination ceases at time zero, we can utilize both the UV rate constant, *k*_*uv*_, and a rate constant, *k*_*v*_, for the ventilation (i.e. *n* ÷ 3600), to produce a decay model for the room space: 

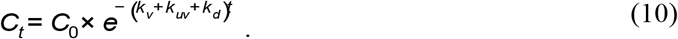

Where; *C*_*0*_ and *C*_*t*_ are the concentrations of viable viral particles in the room space (virions/m^3^) at time zero and *t* seconds respectively; *k*_*v*_ is the ventilation rate constant; *k*_*d*_ is the particulate deposition rate constant (e.g. 0.0014 s^-1^ [19]); and *t* is time in seconds.

### 2.2 Analysis of published data

A search of the relevant scientific literature was undertaken to identify published data relating to the UV irradiation of the three closely related coronaviruses: SARS-CoV-2, the causative agent of COVID-19; SARS-CoV-1, the causative agent of severe acute respiratory syndrome (SARS); and MERS-CoV, the causative agent of middle east respiratory syndrome (MERS).

Because the experimental methods used in the various UV studies varied greatly, as did the level of detail reported, it was necessary to adopt a standardized approach so that valid comparisons could be made. It was therefore decided that, rather than estimating the *Z* value for a nominal log one reduction (i.e. D_90_) as others have done [28], we would instead use the log reduction values and UV doses reported in the various studies to calculate the respective *Z* values using equation 3. In so doing, we were able to utilize the results from studies that would otherwise be excluded because the log reductions achieved were far in excess of one. Where researchers performed experiments using a range of UV doses, we calculated the *Z* value for two UV doses, one near the start of the inactivation process and the other just before the saturation point.

In order to compare the *Z* values for the coronaviruses with those for influenza, we utilized experimental results produced by Heimbuch & Harnish [4] who irradiated coupons of respirator material inoculated with SARS-CoV-1 and MERS-CoV, as well as four strains of influenza A, allowing direct comparisons to be made between the viral species.

### 2.3 Estimating an effective upper-room Z value for aerosolised SARS-CoV-2

In order to evaluate how SARS-CoV-2 might behave in the presence of UV-C when aerosolised, we reviewed the available literature on the subject [25, 28-31] with the aim of estimating a value for the coefficient, *c*_*ur*_, in equation 7, which we then used to estimate the effective upper-room *Z* value, *Z*_*ur*_. In order to reflect the uncertainty associated with this, we compared effective *Z* values for aerosolised coronaviruses reported in the literature with values obtained for SARS-CoV-1 in liquids to obtain the range of possible values for *c*_*ur*_.

### 2.4 Computation of required upper-room UV irradiation flux

Having estimated the value of *Z*_*ur*_ for SARS-CoV-2 from the literature, we then used equations 6 and 8 to estimate the average upper-room irradiation flux that would be required to achieve a 50 - 90% reduction in aerosolised SARS-CoV-2 virions (through the action of the UV-C alone) in a 4.2 × 4.2 × 2.5 m high room space for a range of ventilation rates. These dimensions were chosen because they are typical for an upper-room UVGI installation in which the lamp height is 2.1 m above the floor [15]. In the model we assumed that the air was completely mixed, which meant that according to equation 6, aerosol particles would spend on average 16% of their room residency time in the UV zone.

In addition to computing the required UV flux, we also wanted to know how a standard upper-room UV fitting might perform when challenged by SARS-CoV-2. In accordance with the guidelines stated by First [15], we assumed that the room contained a single 30 W (input) UV fitting capable of delivering an average upper-room flux of 50 μW/cm^2^, and modelled its performance in terms of equivalent ventilation rate using equation 9.

## 3. Results

### 3.1 Analysis of the published literature

The results of the literature search are summarized in Table 1, which shows the UV-C doses applied and log reductions achieved in six studies investigating SARS-CoV-1 and two studies investigating MERS-CoV. Although no studies were found that specifically looked at the inactivation of SARS-CoV-2 using UV-C light, three studies were found that used a combination of UV-A and UV-B light (270-360 nm), together with the photosensitiser, riboflavin, to disinfect SARS-CoV-2 [1, 2] and MERS-CoV [32] in blood products (Table 2). Although these studies did not utilize UV-C light, it was nevertheless decided to report the results of these studies here so that direct comparisons could be made between SARS-CoV-2 and MERS-CoV. The MERS-CoV irradiation study by Bedell et al. [33] is included for completeness, even though the authors did not report the UV dose received by the virus, making it impossible to compute a *Z* value for this study.

**Table 1.**
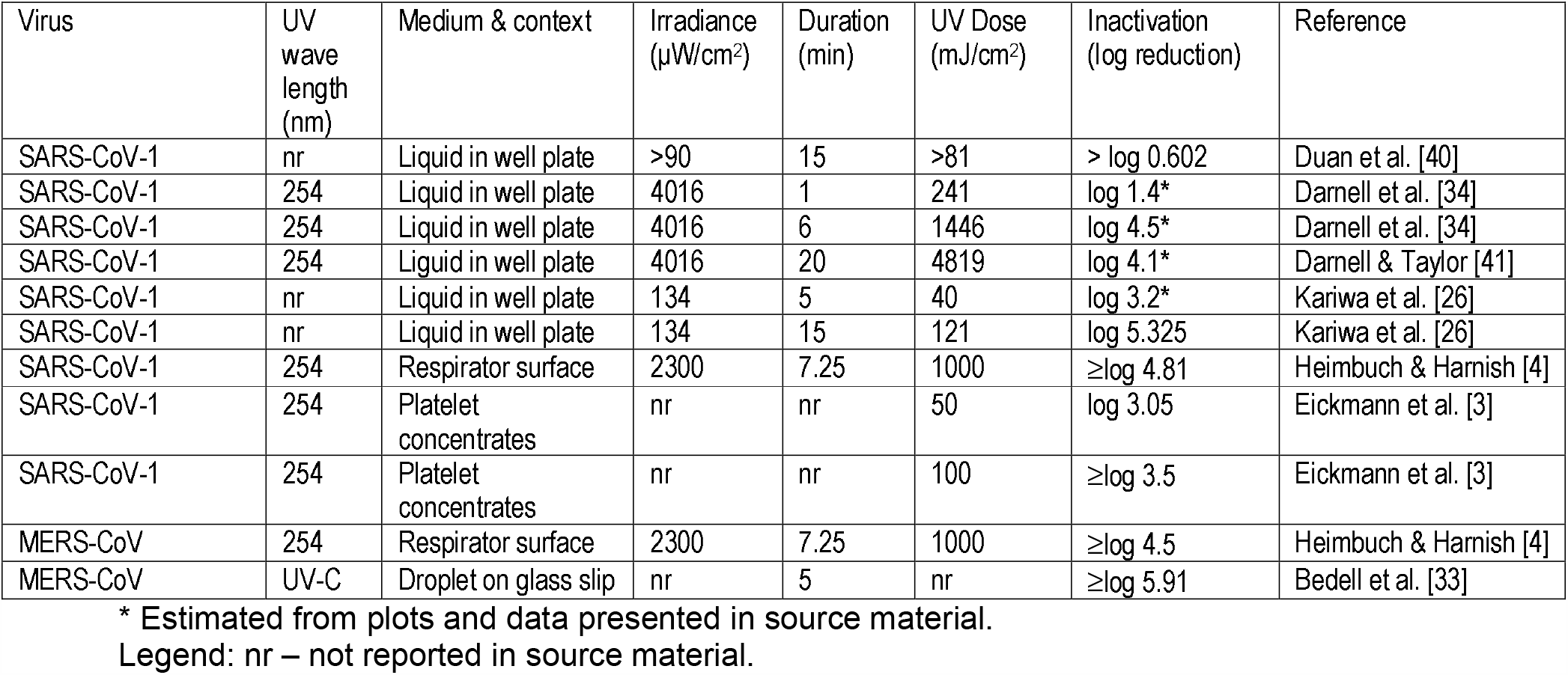
UV-C doses applied and log reductions achieved in various studies relating to the SARS-CoV-1 and MERS-CoV viruses.

| Virus | UV wave length (nm) | Medium & context | Irradiance ($\mu\text{W}/\text{cm}^2$) | Duration (min) | UV Dose ($\text{mJ}/\text{cm}^2$) | Inactivation (log reduction) | Reference |
| --- | --- | --- | --- | --- | --- | --- | --- |
| SARS-CoV-1 | nr | Liquid in well plate | >90 | 15 | >81 | > log 0.602 | Duan et al. [40] |
| SARS-CoV-1 | 254 | Liquid in well plate | 4016 | 1 | 241 | log 1.4* | Darnell et al. [34] |
| SARS-CoV-1 | 254 | Liquid in well plate | 4016 | 6 | 1446 | log 4.5* | Darnell et al. [34] |
| SARS-CoV-1 | 254 | Liquid in well plate | 4016 | 20 | 4819 | log 4.1* | Darnell & Taylor [41] |
| SARS-CoV-1 | nr | Liquid in well plate | 134 | 5 | 40 | log 3.2* | Kariwa et al. [26] |
| SARS-CoV-1 | nr | Liquid in well plate | 134 | 15 | 121 | log 5.325 | Kariwa et al. [26] |
| SARS-CoV-1 | 254 | Respirator surface | 2300 | 7.25 | 1000 | ≥log 4.81 | Heimbuch & Harnish [4] |
| SARS-CoV-1 | 254 | Platelet concentrates | nr | nr | 50 | log 3.05 | Eickmann et al. [3] |
| SARS-CoV-1 | 254 | Platelet concentrates | nr | nr | 100 | ≥log 3.5 | Eickmann et al. [3] |
| MERS-CoV | 254 | Respirator surface | 2300 | 7.25 | 1000 | ≥log 4.5 | Heimbuch & Harnish [4] |
| MERS-CoV | UV-C | Droplet on glass slip | nr | 5 | nr | ≥log 5.91 | Bedell et al. [33] |

\* Estimated from plots and data presented in source material.
Legend: nr – not reported in source material.

**Table 2.** UV-A/B doses applied and log reductions achieved in the various studies relating to the disinfection of SARS-CoV-2 and MERS-CoV in blood products when riboflavin is used.

| Virus | UV wave length (nm) | Medium & context | Irradiance ($\mu\text{W}/\text{cm}^2$) | Duration (min) | UV Dose ($\text{mJ}/\text{mL}$) | Inactivation (log reduction) | Reference |
| --- | --- | --- | --- | --- | --- | --- | --- |
| MERS-CoV | 270-360 | Blood plasma + riboflavin (pooled) | nr | nr | 6240 | ≥log 4.07 | Keil et al. [32] |
| MERS-CoV | 270-360 | Blood plasma + riboflavin (single donor) | nr | nr | 6240 | ≥log 4.42 | Keil et al. [32] |
| SARS-CoV-2 | 270-360 | Blood plasma + riboflavin | nr | 4 | 1872 | ≥log 2.61 | Ragan et al. [1] |
| SARS-CoV-2 | 270-360 | Blood plasma + riboflavin | nr | 4 | 3744 | ≥log 4.72 | Ragan et al. [1] |
| SARS-CoV-2 | 270-360 | Blood plasma + riboflavin | nr | nr | 6240 | ≥log 3.4 | Keil et al. [2] |
| SARS-CoV-2 | 270-360 | Platelets + riboflavin | nr | nr | 6240 | ≥log 4.53 | Keil et al. [2] |

Legend: nr – not reported in source material.

The computed *Z* values for the respective experiments are shown in Tables 3 (UV-C) and 4 (UV-A/B plus riboflavin). From these it can be seen that the *Z* values for the MERS-CoV virus were similar in magnitude to those for both SARS-CoV-1 (UV-C) and SARS-CoV-2 (UV-A/B). With UV-C irradiation the mean *Z* value for SARS-CoV-1 was 0.00489 (SD = 0.00611) m^2^/J, whereas that for MERS-CoV was 0.00104 m^2^/J. Likewise, for UV-A/B plus riboflavin the corresponding *Z* values were 0.00020 (SD = 0.00009) m^2^/J and 0.00016 m^2^/J for SARS-CoV-2 and MERS-CoV respectively.

**Table 3.** Calculated *Z* values for the UV-C irradiation experiments.

| Virus | UV Dose (mJ/cm<sup>2</sup>) | Inactivation (log reduction) | UV susceptibility constant, Z (m<sup>2</sup>/J) | Reference |
| --- | --- | --- | --- | --- |
| SARS-CoV-1 | >81 | > log 0.602 | 0.00171 | Duan et al. [40] |
| SARS-CoV-1 | 241 | log 1.4* | 0.00134* | Darnell et al. [34] |
| SARS-CoV-1 | 1446 | log 4.5* | 0.00072* | Darnell et al. [34] |
| SARS-CoV-1 | 4819 | log 4.1* | 0.00020* | Darnell & Taylor [41] |
| SARS-CoV-1 | 40 | log 3.2* | 0.01833* | Kariwa et al. [26] |
| SARS-CoV-1 | 121 | log 5.325 | 0.01017 | Kariwa et al. [26] |
| SARS-CoV-1 | 1000 | ≥log 4.81 | 0.00111 | Heimbuch & Harnish [4] |
| SARS-CoV-1 | 50 | log 3.05 | 0.01405 | Eickmann et al. [3] |
| SARS-CoV-1 | 100 | ≥log 3.5 | 0.00806 | Eickmann et al. [3] |
| MERS-CoV | 1000 | ≥log 4.5 | 0.00104 | Heimbuch & Harnish [4] |

\* Estimated from plots and data presented in source material.

**Table 4.** Calculated *Z* values for the UV-A/B irradiation plus riboflavin experiments.

| Virus | UV Dose (mJ/mL) | Inactivation (log reduction) | UV susceptibility constant, Z (m<sup>2</sup>/J) | Reference |
| --- | --- | --- | --- | --- |
| MERS-CoV | 6240 | ≥log 4.07 | 0.00015 | Keil et al. [32] |
| MERS-CoV | 6240 | ≥log 4.42 | 0.00016 | Keil et al. [32] |
| SARS-CoV-2 | 1872 | ≥log 2.61 | 0.00032 | Ragan et al. [1] |
| SARS-CoV-2 | 3744 | ≥log 4.72 | 0.00029 | Ragan et al. [1] |
| SARS-CoV-2 | 6240 | ≥log 3.4 | 0.00013 | Keil et al. [2] |
| SARS-CoV-2 | 6240 | ≥log 4.53 | 0.00017 | Keil et al. [2] |

The calculated *Z* values for influenza UV-C irradiation experiments undertaken by Heimbuch & Harnish [4] are presented in Table 5. These experiments, which were carried out using inoculated coupons of respirator material, revealed that in this context the *Z* values for the various influenza A strains were of the same order of magnitude as those for SARS-CoV-1 and MERS-CoV.

**Table 5.**
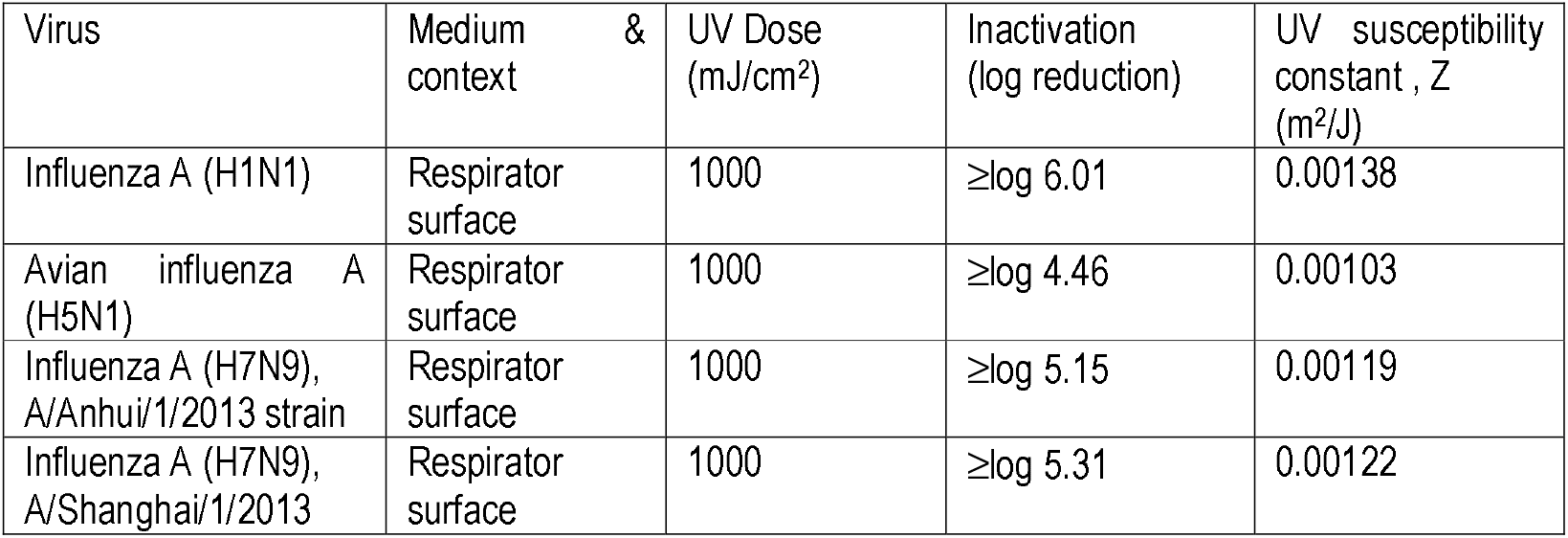
Calculated *Z* values for the UV-C irradiation experiments for different strains of influenza A tested by Heimbuch & Harnish [4].

### 3.2 Effective upper-room Z values for aerosolised SARS-CoV-2

A review of the literature revealed that relatively few experimental studies have been performed involving the UV irradiation of aerosolised viruses, with only one undertaken on a coronavirus [29]. A summary of the findings of several key studies are presented in Table 6, which reveals that most viral species appear to be relatively easy to disinfect when suspended in droplets in the air. In particular, aerosolised viruses appear to be more vulnerable to UV damage than when they are suspended in a liquid or on a substrate. For example, for the 24 irradiation experiments involving adenoviruses suspended in liquid, reported by Kowalski [28], the average *Z* value was 0.00586 m^2^/J, which is an order of magnitude less than the values of 0.0546 and 0.0390 m^2^/J for aerosolised adenoviruses, attributed to Jensen [31] and Walker and Ko [29] respectively. Regarding coronaviruses, Walker and Ko [29] also performed experiments on aerosolised murine (mouse) hepatitis virus (MHV) coronavirus in a single pass test rig. This revealed a *Z* value of 0.377 ± 0.119 m^2^/J for this virus, which is several orders of magnitude greater than the values obtained above for SARS-CoV-1 and MERS-CoV in liquids and on surfaces (Table 3). Although we are comparing different species of coronavirus here, evidence from Bedell et al. [33], who irradiated MHV coronavirus and MERS-CoV in Petri dishes, suggests that it is nonetheless valid. They found that 5 minutes exposed to a UV-C light source resulted in a 2.71 log reduction for the MHV coronavirus, whereas the same exposure resulted in a 5.91 log reduction for MERS-CoV. This suggests that MHV coronavirus is actually more resistant to UV damage than MERS-CoV, and as such, supports Walker and Ko’s [29] conclusion that coronaviruses are much easier to inactivate in the air compared with on surfaces and in liquids.

**Table 6.** Summary of reported effective Z values for single-pass UV irradiation experiments performed on aerosolised viruses in air.

| Researchers | Virus | Effective Z value (m<sup>2</sup>/J) | Reporter |
| --- | --- | --- | --- |
| Jensen [31] | Adenovirus | 0.0546 | Kowalski et al. [30] |
| Jensen [31] | Coxsackie B-1 | 0.1108 | Kowalski et al. [30] |
| Jensen [31] | Influenza A | 0.1187 | Kowalski et al. [30] |
| Jensen [31] | Sindbis virus | 0.1040 | Kowalski [28] |
| Jensen [31] | Vaccinia virus | 0.1528 | Kowalski et al. [30] |
| Walker & Ko [29] | Adenovirus | 0.0390 | Walker & Ko [29] |
| Walker & Ko [29] | MHV coronavirus | 0.3770 | Walker & Ko [29] |
| McDevitt et al. [25] | Influenza A | 0.2700 | McDevitt et al. [25] |
| McDevitt et al. [42] | Vaccinia virus | 2.5400 | McDevitt et al. [42] |

Comparing the computed *Z* values for UV irradiation experiments on coronaviruses conducted in air (0.37700 m^2^/J [29]) with equivalents in liquid (0.00134 m^2^/J) [34]; 0.01833 m^2^/J [26], it would appear that irradiating the coronavirus in liquid requires a UV dose that is in the region 20 - 281 times higher than that required when the virus is suspended in air. From this we estimated that the value of the adjustment coefficient *c*_*ur*_ would be in a range 0.05 – 0.0036.

### 3.3 Upper-room UVGI simulation results

Because the *Z* value for aerosolised MHV coronavirus appeared to be indicative of how airborne SARS-CoV-2 might behave in a UV-C field, a decision was made to use Walker and Ko’s mean *Z* value of 0.377 m^2^/J to evaluate the expected performance of an upper-room UVGI installation. Notwithstanding this, because of the uncertainty associated with how aerosolised SARS-CoV-2 might behave in reality, we also modelled a worst-case scenario in which *Z*_*ur*_ was 0.0377 m^2^/J.

Table 7 presents the results of the room analysis using these two values for *Z*_*ur*_, for a range of ventilation rates. From this it can be seen that there is a direct inverse relationship between particle residence time in the UV field, *t*_*uv*_, and the required irradiation flux, *E*_*r*_, as predicted by equation 8. This means that for any given *Z* value, the value of *E*_*r*_ will double as the room ventilation rate doubles. The table also reveals that there is a direct inverse relationship between *Z*_*ur*_ and *E*_*r*_. From the calculated values in this table it can be seen that if *Z*_*ur*_, = 0.377 m^2^/J, then with an average UV flux of just 10 μW/cm^2^ it should be possible to achieve >90% inactivation of SARS-CoV-2, even at a ventilation rate of 8 AC/h. However, if in reality, *Z*_*ur*_, is 0.0377 m^2^/J, then all the calculated fluxes would have to increase by a factor of ten to achieve the same results. Given that accepted guidelines [15] recommend for a room 2.5 m high, one 30 W (input) UV lamp per 18.58 m^2^ of floor area, which will produce an average flux in the region 50 μW/cm^2^, this means that even under this worse-case scenario it should still be possible to achieve disinfection rates >90% for all but the highest ventilation rates.

**Table 7.** Predicted average upper-room UV irradiance fluxes required to achieve 50%, 70% and 90% inactivation for SARS-CoV-2 assuming a range of *Z*_*ur*_ values and ventilation rates. (Assuming *Z*_*ur*_ = 0.377 or 0.0377 m^2^/J)

| Ventilation rate (AC/h) | Average particle residence time in UV field. (mins.) | UV susceptibility constant, $Z_{ur}$ (m<sup>2</sup>/J) | Average irradiance required for 50% inactivation (μW/cm<sup>2</sup>) | Average irradiance required for 70% inactivation (μW/cm<sup>2</sup>) | Average irradiance required for 90% inactivation (μW/cm<sup>2</sup>) |
| --- | --- | --- | --- | --- | --- |
| 1 | 9.6 | 0.3770 | 0.319 | 0.554 | 1.060 |
| 2 | 4.8 | 0.3770 | 0.638 | 1.109 | 2.121 |
| 4 | 2.4 | 0.3770 | 1.277 | 2.218 | 4.241 |
| 6 | 1.6 | 0.3770 | 1.915 | 3.327 | 6.362 |
| 8 | 1.2 | 0.3770 | 2.554 | 4.436 | 8.482 |
| 1 | 9.6 | 0.0377 | 3.192 | 5.544 | 10.604 |
| 2 | 4.8 | 0.0377 | 6.384 | 11.088 | 21.207 |
| 4 | 2.4 | 0.0377 | 12.768 | 22.177 | 42.414 |
| 6 | 1.6 | 0.0377 | 19.152 | 33.266 | 63.621 |
| 8 | 1.2 | 0.0377 | 25.536 | 44.355 | 84.829 |

When we fixed the UV flux at an average of 50 μW/cm^2^, we found that for *Z*_*ur*_, = 0.377 m^2^/J the upper-room UVGI installation produced an equivalent air change rate of 108.6 AC/h, whereas if *Z*_*ur*_, = 0.0377 m^2^/J this fell to 10.9 AC/h. These values were constant and unaffected by the actual room ventilation rate.

## 4. Discussion

Although the impact of UV-C on SARS-CoV-2 has not yet been experimentally investigated for aerosols, the results of our analysis suggest that it is highly lightly that the UV-C *Z* value for this virus will be similar in magnitude to that for SARS-CoV-1 and MERS-CoV. This is because MERS-CoV behaves very similarly to SARS-CoV-1 when exposed to UV-C light (Table 3) and also very similar to SARS-CoV-2 when exposed to UV-A/B and riboflavin (Table 4). As such, there are grounds for believing that the UV-C susceptibility constant, *Z*, for SARS-CoV-2 will be similar to those of the other two coronaviruses.

One problem often encountered when comparing UV irradiation results from disparate researchers is that experimenters often utilize different methodologies to evaluate log reductions in microbial species, with varying doses of UV administered. In particular, the type of substrate or media used can greatly influence the outcome of the experiment. This is because the substrate or media can absorb the UV photons and shield the virus. Given this, it is important to compare like with like, if this is possible. For this reason we included the results of Heimbuch and Harnish [4] in Tables 3 and 5, because they performed the same irradiation experiment on SARS-CoV-1 and MERS-CoV, as well as on four strains of influenza A, thus allowing direct comparisons to be made. From Tables 3 and 5 it can be seen that the *Z* values for the influenza strains are of a similar order of magnitude as those for the coronaviruses, implying that in this context SARS-CoV-1 and MERS-CoV were about as difficult to inactivate as influenza A. This is a surprising finding, because others have suggested that the UV dose required to disinfect SARS-CoV-2 might be much higher than that required to disinfect influenza A [5]. Indeed, in a summary collated from hundreds of published studies by Kowalski [28], the Z values for influenza A in water were reported as being in the range 0.04800 - 0.13810 m^2^/J, much higher than the values achieved by Heimbuch and Harnish [4]. As such, this suggests that the substrate or medium in which microbe is irradiated plays an important role in influencing the magnitude of the *Z* value achieved. Indeed, it is well known in other contexts that UV-C light can be attenuated as it passes through liquids [35]. When UV light passes through a suspension of particles in water, its intensity is reduced due to both scattering and absorption of the light [36]. Absorption occurs because the light beam interacts with atoms and molecules in the liquid to raise their energy level, with the result that energy is lost from the beam, whereas scattering occurs when particulates in the fluid interfere with the UV light making it more diffuse [35]. Particulates can also shield microbes from UV light. This means that UV inactivation of microbial suspensions in liquid films >1.2 mm can be greatly inhibited, due to the low penetration depth of UV light through concentrated suspensions [37]. Consequently, when interpreting the Z values for SARS-CoV-1 and MERS-CoV in Table 3, it is important to view them as being strictly contextual.

With regard to UV irradiation of aerosolised viruses, very few published experimental studies exist, with only one specifically relating to a coronavirus [29]. As a result there is a paucity of good quality data relating to UV-C irradiation of SARS-CoV-1, SARS-CoV-2 and MERS-CoV in the air. Consequently, we had to establish whether or not Walker and Ko’s [29] published *Z* value of 0.377 m^2^/J was valid for SARS-CoV-2 in air. Comparison with the Z values presented in Table 3 reveals that this value is several orders of magnitude greater than those achieve when coronaviruses are irradiated in liquid or on equipment substrates. This however, is to be expected given that liquids attenuate UV penetration [35]. Also the finding appears to be broadly in keeping with the behaviour of adenoviruses when irradiated in air and in liquid. Furthermore, because Bedell et al. [33] found MERS-CoV to be more susceptible to UV-C damage than MHV coronavirus, this strongly supports the use of Walker and Ko’s [29] *Z* value for MHV coronavirus as a valid surrogate for SARS-CoV-2 in air. Having said this, because the UV susceptibility of the target microbe is crucial to the performance of any upper-room UVGI installation, Walker and Ko’s *Z* value for coronaviruses should be treated with caution, as it may in time turn out to be incorrect. For this reason, when we assessed the performance of the upper-room UVGI in our hypothetical room, we used both 0.377 and 0.0377 m^2^/J in our simulations. In so doing, we effectively modelled both the expected and worst-case scenarios.

The results for the expected and worst-case scenarios in Table 7, strongly suggest that upper-room UVGI, if applied correctly, should be effective at disinfecting SARS-CoV-2 virions suspended in respiratory droplets in the air. This finding is of course is very much dependent on the surrogate *Z*_*ur*_ value being truly representative for SARS-CoV-2. With respect to this, one limitation of our study is that we did not distinguish between the *Z* values achieved using a single-pass test rig, such as that used by Walker and Ko [29], and those achieved in real-life by an upper-room UVGI system. With the latter, because the irradiation process is fragmented, compared with a single-pass system, it is thought that higher UV doses might be required to achieve equivalent levels of inactivation [12, 27]. However, while this specifically applies to aerosolised bacteria that can rapidly repair UV damage when the irradiation process becomes fragmented [38], it is not known to what extent this applies to viruses, which are not metabolically active, although it is known that through photoreactivation viruses can repair UV damage [39].

One great advantage of upper-room UVGI is that it can be retrospectively fitted into buildings provided that the floor to ceiling height is large enough to ensure that the UV field does not impinge on room occupants [15]. By installing such as system it is possible to effectively ‘turbo-charge’ the efficacy of the ventilation system. Indeed, our analysis suggests that it is possible to achieve >100 equivalent AC/h by installing upper-room UVGI. Using equation 9, we can calculate the UV rate constant, *k*_*uv*_, which can be thought of as the equivalent air change rate per second. Once known, this in turn can be used, together with the ventilation and particulate deposition rate constants, *k*_*v*_, and *k*_*d*_, in equation 10, to compute the concentration of viral partials in the room space at any point in time.

## 5. Conclusion

We have been able to demonstrate that the UV-C susceptibility constant, *Z*, for SARS-CoV-2 is likely to be similar to that exhibited by the SARS-CoV-1 and MERS-CoV viruses. Furthermore, we have found evidence suggesting that SARS-CoV-2 when suspended in air is reasonably easy to inactivate using UV light at 254 nm. As such, this suggests that upper-room UVGI may have great potential as an intervention to inhibit the transmission of COVID-19 in buildings, especially in situations where achieving high ventilation rates might otherwise be impractical.

## Data Availability

All the data used in the study is in the public domain and has been published in cited journal articles and books.

